# Eating disorder phenotypes differ in their genetic relationships with impulsivity facets

**DOI:** 10.64898/2026.11.23.26367764

**Authors:** Laura Vilar-Ribó, Ryder Digiulio, Sevim Bianchi, Maia Choi, Daniel E Gustavson, Sandra Sanchez-Roige

## Abstract

**Background:** Eating disorders are clinically heterogeneous and characterized by distinct patterns of control over food intake. Anorexia nervosa (**AN**) is characterized by excessive dietary restraint, whereas binge-eating (**BE**) is a transdiagnostic behavior defined by loss of control over eating. Understanding the relationship between eating disorders and impulsivity may provide insights into their underlying mechanisms and identify potential targets for prevention and treatment.

**Methods:** We used genome-wide association study data (N=82,707-1,268,116) and genomic structural equation modeling to examine genetic relationships between eating disorder phenotypes (AN, AN_Restrictive_, AN_Binge-Purging_, and BE) and impulsivity facets (five UPPS-P, three BIS-11, and delay discounting). We examined whether these relationships persisted after conditioning on the genetic liability for substance use disorders (**SUDs**) and body mass index (**BMI**).

**Results:** AN and its subtypes showed negative genetic correlations with delay discounting, the magnitude of this correlation being stronger for AN_Restrictive_. AN_Restrictive_ also showed negative genetic correlations with sensation seeking and common impulsivity, whereas AN_Binge-Purging_ showed a positive genetic correlation with urgency-specific impulsivity. In contrast, BE showed positive genetic correlations with both common and urgency-specific impulsivity. Several associations remained significant after controlling for SUDs and BMI, including the negative association between AN with delay discounting, the negative associations with AN and AN_Restrictive_ with sensation seeking, and the positive association between BE and urgency- specific impulsivity.

**Conclusion:** These findings reveal complex patterns of genetic overlap between eating disorder phenotypes and impulsivity dimensions. Understanding these phenotype-specific relationships may help explain clinical heterogeneity and inform the development of more targeted prevention and treatment strategies.

## INTRODUCTION

Eating disorders are prevalent, affecting approximately 8% of women and 4% of men over their lifetime (Harvard, 2020). Eating disorders are heterogeneous, and characterized by distinct patterns of control over food intake. Anorexia nervosa (**AN**) is characterized by persistent restriction of energy intake, fear of weight gain or behaviors that interfere with weight gain, and disturbances in body image. AN includes two subtypes that differ in their patterns of eating behavior: individuals with the restrictive subtype (**AN_Restrictive_**) primarily engage in dietary restriction (Puckett et al., 2021), whereas those with the binge-eating/purging subtype (**AN_Binge-Purging_**) also engage in recurrent binge-eating (**BE**) and/or purging behaviors (Uniacke et al., 2020). Both subtypes are typically, although not always, accompanied by low body mass index (**BMI**) (De Young et al., 2013). BE is a transdiagnostic behavior shared across some eating disorders, including bulimia nervosa and AN_Binge-Purging_, and is characterized by episodes of consuming large amounts of food in discrete periods of time accompanied by a sense of loss of control (Berkman et al., 2015; Giel et al., 2022). Individuals who engage in BE often have high BMI, although it occurs across the weight spectrum (McCuen-Wurst et al., 2018). These distinct patterns of eating behaviors have been linked to differences in behavioral control, with dietary restriction associated with excessive self-control and BE characterized by episodes of loss of control. Thus, food restriction and BE capture partly opposing dimensions of control over eating, while also co-occurring within some individuals and diagnoses, or oscillating over time as individuals transition between diagnoses (Eddy et al., 2008; Schaumberg et al., 2019). A deeper understanding of the relationship between eating disorders and self-control mechanisms (i.e., impulsivity) may offer novel avenues for prevention and treatment strategies (Bardone-Cone et al., 2016; Boswell et al., 2023; İnce et al., 2021; Testa et al., 2022).

Individuals with AN often show *low* levels of impulsivity and excessive control over food intake, even when such behavior is life-threatening (Butler & Montgomery, 2005; Fessler, 2002; Howard et al., 2020; Mallorquí-Bagué et al., 2020; Meneguzzo et al., 2021; Steinglass et al., 2012; Steward et al., 2017), whereas individuals who engage in BE often show *high* levels of impulsivity (Carr et al., 2021). Individuals with AN_Binge-Purging_ show co-occurring low and high levels of impulsivity (Lavender & Mitchell, 2015; Meneguzzo et al., 2021). However, impulsivity is multifaceted (Dick et al., 2010; Evenden, 1999; Griffin et al., 2018; Levitt et al., 2020; Meda et al., 2009; Miller & Gizer, 2024) and encompasses different tendencies that may contribute differently to eating disorder phenotypes, including delay discounting, or the preference for delayed rewards, such as forgoing food and overriding hunger cues in order to maintain a low body weight (Decker et al., 2015); risk-taking, such as engaging in dangerous eating-related behaviors despite potential harm (e.g., purging, laxative misuse) (Stein et al., 2020); and negative urgency, or the tendency to act impulsively to alleviate negative emotions or distress, potentially contributing to BE (Bardone-Cone et al., 2016). These observations suggest that different impulsivity facets may show distinct, and potentially opposing, relationships with eating disorder phenotypes.

Both eating disorders and impulsivity are highly heritable traits (Bulik et al., 2016; Sanchez-Roige et al., 2018, 2019, 2023; Termorshuizen et al., 2026; Thorpe et al., 2024). In a previous study, we found negative genetic correlations between AN and specific facets of impulsivity, including delay-discounting and lack of perseverance (Bianchi et al., 2025).

However, these analyses could not establish whether these genetic relationships differed across eating disorder phenotypes because genome-wide association studies (**GWAS**) of AN subtypes and BE were not yet available.

With the availability of recent large-scale GWAS from the Psychiatric Genomics Consortium Eating Disorders (**PGC-ED**) Workgroup (Termorshuizen et al., 2026), we extended our prior work to examine shared and distinct genetic relationships between four eating disorder phenotypes (AN, AN_Restrictive_, AN_Binge-Purging_, and BE) and nine impulsivity facets. To capture the multidimensional nature of impulsivity, we used GWAS summary statistics derived from the Impulsive Behavior Scale (**UPPS-P**; Cyders et al., 2014; Whiteside & Lynam, 2001), the Barratt Impulsiveness scale (**BIS**; Patton et al., 1995), and the Monetary Choice Questionnaire (**MCQ**; Kirby et al., 1999). We first estimated pairwise genetic correlations between eating disorder phenotypes and impulsivity facets. We then examined whether these relationships persisted after accounting for genetic liability to substance use disorders (**SUDs**) and body mass index (**BMI**). Impulsivity facets share positive genetic correlations with SUDs (Mitchell, 2019; Sanchez-Roige et al., 2018, 2023; Vilar-Ribó et al., 2025), which also share genetic overlap with eating disorders (Bahji et al., 2019; Devoe et al., 2021; Mellentin, Mejldal, et al., 2022; Mellentin, Nielsen, et al., 2022; Munn-Chernoff et al., 2021). Similarly, AN and BE show strong but opposing genetic relationships with BMI (Toppino et al., 2022). We therefore used multivariable genomic regression to evaluate whether the observed genetic relationships between eating disorder phenotypes and impulsivity facets were independent of shared genetic liability to SUDs and BMI.

Based on previous clinical and genetic findings, we tested three hypotheses: i) AN would show negative genetic correlations with delay discounting and perseverance, particularly for AN_Restrictive_; ii) AN_Binge-Purging_ would negative genetic correlations with delay discounting but positive genetic correlations with urgency-related facets, consistent with the co-occurrence of persistent dietary restriction and episodes of loss of control over eating; and iii) BE would show positive genetic correlations with impulsivity, particularly with urgency-related facets.

## METHODS

### Genome-wide association studies

*Overview.* **Table 1** provides an overview of the GWAS datasets used. We used summary statistics from published GWAS of individuals genetically similar to European-like reference populations (National Academies of Sciences, Engineering, and Medicine, 2023). Our study was limited to this group because GWAS for these phenotypes are not currently available at scale for other groups.

**Table 1.** Sample sizes and references for genome-wide association studies. All traits were coded such that higher values equal higher endorsement of that trait or increased likelihood of that disorder. Samples were limited to individuals of European-like ancestry. Sample sizes for binary traits are reported as *N* effective. *BE case samples were not ascertained for AN.

| Trait | Abbreviation | Sample Size | Sample Type | Reference |
|---|---|---|---|---|
| **Anorexia nervosa** | AN | 94,741 | Cases (24,145) Controls (1,243,971) | (Watson et al., 2019) |
| **Anorexia nervosa restrictive** | AN<sub>Restrictive</sub> | 10,076 | Cases (2,524) Controls (1,243,971) | (Termorshuizen et al., 2025) |
| **Anorexia nervosa binge-purge** | AN<sub>BingePurging</sub> | 20,892 | Cases (5,245) Controls (1,243,971) | (Termorshuizen et al., 2025) |
| **Binge eating\*** | BE | 125,541 | Cases (15,179) Controls (1,227,436) | (Termorshuizen et al., 2025) |
| **UPPS- P: Negative Urgency** | NU | 132,559 | Continuous | (Sanchez-Roige et al., 2023) |
| **UPPS- P: Positive Urgency** | PU | 132,132 | Continuous | (Sanchez-Roige et al., 2023) |
| **UPPS- P: Lack of premeditation** | Premed | 132,667 | Continuous | (Sanchez-Roige et al., 2023) |
| **UPPS- P: Lack of perseverance** | Perseverance | 133,517 | Continuous | (Sanchez-Roige et al., 2023) |
| **UPPS- P: Sensation Seeking** | SS | 132,395 | Continuous | (Sanchez-Roige et al., 2023) |
| **BIS: Attentional** | Attention | 124,739 | Continuous | (Sanchez-Roige et al., 2023) |
| **BIS: Motor** | Motor | 124,104 | Continuous | (Sanchez-Roige et al., 2023) |
| **BIS: Nonplanning** | Nonplan | 123,509 | Continuous | (Sanchez-Roige et al., 2023) |
| **Body mass index** | BMI | 694,649 | Continuous | (Pulit et al., 2019) |
| **Delay Discounting** | DD | 134,935 | Continuous | (Thorpe et al., 2026) |
| **Problematic alcohol use** | PAU | 903,147 | Cases (263,224) Controls (639,923) | (Zhou et al., 2023) |
| **Cannabis use disorder** | CUD | 161,053 | Cases (42,281) Controls (843,744) | (Levey et al., 2023) |
| **Opioid use disorder** | OUD | 36,799 | Cases (10,544) Controls (72,163) | (Zhou et al., 2020) |
| **Tobacco use disorder** | TUD | 532,367 | Cases (174,021) Controls (565,874) | (Toikumo et al., 2024) |

*Eating disorders.* We used summary statistics from the most recent publicly available GWAS of eating disorder phenotypes by the PGC-ED, including AN, AN_Restrictive_, AN_Binge-Purging_, BE (Termorshuizen et al., 2026). The clinical sample for AN included 24,223 cases and 1,243,971 controls. Among AN cases with available subtype information, 2,524 were classified as AN_Restrictive_ and 5,245 as AN_Binge-Purging_. For BE, we used GWAS summary statistics from studies not ascertained for AN, including 32,029 cases, and 1,227,436 controls. We selected this subset to minimize the contribution of AN-ascertained samples. For more information about individual phenotypes, refer to Termorshuizen et al. (2026).

*Impulsivity.* We used summary statistics from our latest GWAS of impulsivity based on a sample of up to 133,517 research-consented participants from the 23andMe Research Institute (Sanchez-Roige et al., 2019, 2023). These comprised five UPPS-P Impulsive Behavior Scale facets (Cyders et al., 2014; Whiteside & Lynam, 2001), including lack of premeditation, lack of perseverance, positive urgency, negative urgency, and sensation seeking, and three BIS-11 scale facets, including attentional, motor, and non-planning impulsivity (Patton et al., 1995).

These datasets have been extensively described elsewhere Sanchez-Roige et al. (2019, 2023). *Delay discounting.* We used summary statistics from our latest GWAS of delay discounting (Thorpe et al., 2024), which included 134,935 research-consented participants from the 23andMe Research Institute. Participants completed the 27-item Monetary Choice Questionnaire (Kirby et al., 1999). More information about this dataset can be found in Thorpe et al. (2024), Sanchez-Roige et al. (2018).

*Substance use disorders*. We used summary statistics from four SUDs GWAS: problematic alcohol use (**PAU**; N = 903,147)(Zhou et al., 2023), tobacco use disorder (**TUD**; N = 495,471; (Toikumo et al., 2024), cannabis use disorder (**CUD**; N = 886,025) (Levey et al., 2023) and opioid use disorder (**OUD**; N = 302,585) (Kember et al., 2022).

*BMI*. We used summary statistics from the most recent publicly available GWAS of BMI (N = 694,649;(Pulit et al., 2019).

### Data analyses

All analyses were conducted in *R* (Version 4.1.1; R CoreTeam, 2022). We used the GenomicSEM package version 0.0.4 (Grotzinger et al., 2019), which applies SEM methods to GWAS summary statistics. Genomic SEM leverages Linkage Disequilibrium Score Regression (**LDSC**) to generate a genetic correlation matrix between traits from GWAS summary statistics (Bulik-Sullivan et al., 2015). Genomic SEM accounts for sample overlap by estimating a sampling covariance matrix that captures the precision of the estimates as well as the extent to which the sampling dependencies of the estimates are associated (Grotzinger et al., 2019).

Structural equation models were fit to the data using GenomicSEM, which draws on functionality from the *lavaan R* package (Rosseel, 2012). We used European reference panels from 1000G, and the default diagonally weighted least squares (**DWLS**) estimation method in these analyses. We determined the model fit based on the comparative fit index (**CFI**) and the standardized root mean square residual (**SRMR**). Good-fitting models are expected to have CFI higher than .95 (.90 for acceptable fit) and SRMR smaller than .08 (Hu & Bentler, 1998).

Significance of individual parameter estimates was established with 95% confidence intervals (**CIs**) and FDR corrected *p* value to account for multiple testing.

### Model-fitting approach

In order to examine the relationships of each eating disorder subtypes and impulsivity facets, we modelled each eating disorder GWAS as individual traits. We used a latent factor structure of impulsivity, as in our prior work (Bianchi et al., 2025; Gustavson et al., 2020, 2024). This model included a “common impulsivity” factor that captures shared variance across all three BIS subscales and the UPPS-P lack of premeditation, negative urgency, and positive urgency subscales, as well as an “urgency-specific” impulsivity latent factor capturing additional shared variance among the UPPS-P positive and negative urgency subscales only. Although some impulsivity models include perseverance and sensation seeking, our prior work showed that they are genetically distinguishable and not strong contributors to common impulsivity factors (Mallard et al., 2026; Sanchez-Roige et al., 2019, 2023). Therefore, we modeled lack or perseverance, sensation seeking, and delay discounting as separate indicators (Gustavson et al., 2020, 2024; Vilar-Ribó et al., 2025). Next, we fit a SUDs factor, which replicated a prior addiction risk factor model (Hatoum et al., 2022, 2023), replacing problematic tobacco use for TUD (Toikumo et al., 2024), which was not available at the time of their analyses (Vilar-Ribó et al., 2025). Finally, we constructed a model in which the five impulsivity factors, SUDs factor, eating disorder phenotypes (AN, AN_Binge-Purging_, AN_Restrictive_, BE), and BMI were fitted simultaneously.

We fit two versions of this model: (1) a correlated factors model where we estimated the genetic correlations between eating disorder traits and all impulsivity factors, SUDs, BMI, and (2) a multiple regression model, where eating disorder traits were regressed on impulsivity, SUDs, and BMI, to estimate the genetic associations between each eating disorder phenotype and impulsivity factor after accounting for the shared genetic relationship across impulsivity, SUDs and BMI.

## Data availability

GWAS summary statistics for AN, AN_Binge-Purging_, AN_Restrictive_, BE, and the SUD traits are publicly available. The full GWAS summary statistics for the 23andMe discovery data set will be made available through 23andMe to qualified researchers under an agreement with 23andMe that protects the privacy of the 23andMe participants. Please visit https://research.23andme.com/collaborate/#dataset-access/ for more information and to apply to access the data. The *R* data files containing the genomic SEM matrices for all analyses are displayed at the following link: https://osf.io/6jys3/. This allows for replication and analyses of competing models without obtaining the source data.

## RESULTS

### Genetic correlations among eating disorders, impulsivity, SUDs and BMI phenotypes

Before fitting a Genomic SEM model, we generated a genetic correlation matrix between all study variables. **Figure 1a** shows the genetic correlation (*r_g_*) matrix among eating disorder phenotypes. As expected, genetic correlations among AN and its subtypes were high, ranging from *r_g_ =* 0.61 ± 0.13 to *r_g_ =* 0.87 ± 0.10. Genetic correlations between AN subtypes and BE were much lower (*r_gAN-BE_ =* 0.24 ± 0.05 and *r_gANBingePurging-BE_ =* 0.21 ± 0.08) or not significant (*r_gANRestrictive-BE_ =* -0.01 ± 0.12). **Figure 1b** shows genetic correlations between eating disorder phenotypes, impulsivity facets, SUDs and BMI phenotypes, revealing distinct patterns of genetic overlap across eating disorder phenotypes.

**Figure 1.**
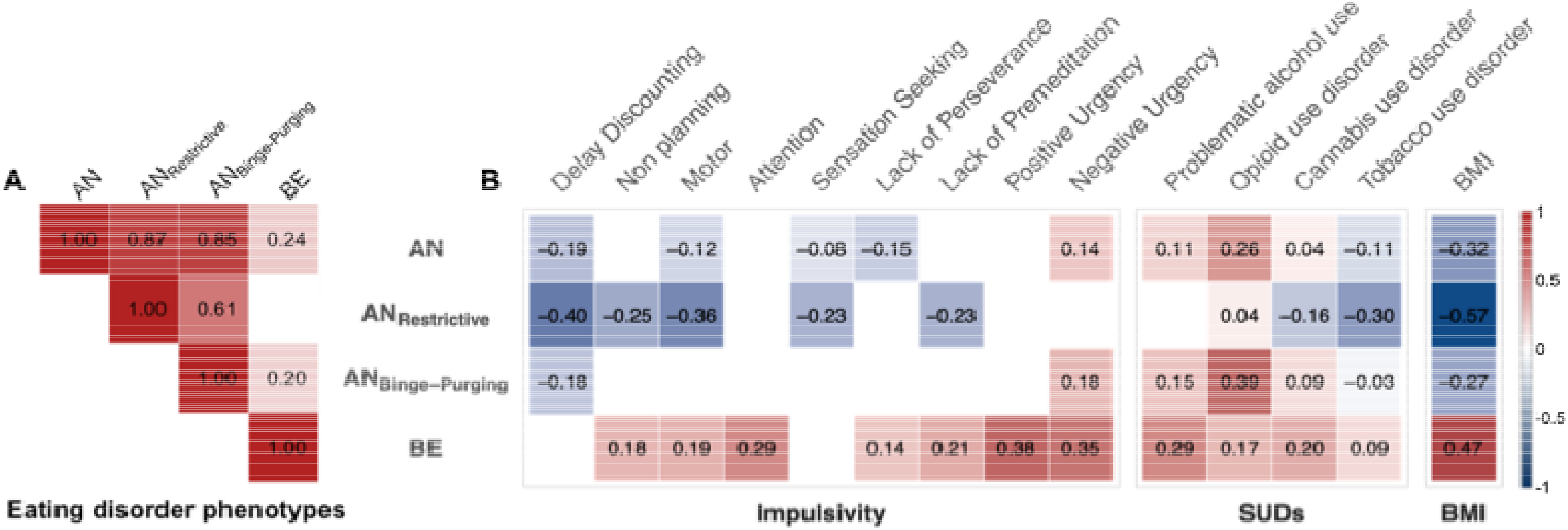
**Genetic correlations (*r_g_*) between study variables**. **(A)** Pairwise *r_g_* between eating disorder phenotypes: AN, anorexia nervosa; AN_Restrictive_, anorexia nervosa restrictive; AN_Binge-Purging_, anorexia nervosa binge-purging; BE, binge-eating. **(B)** *r_g_* between eating disorder phenotypes and impulsivity, substance use disorders (**SUDs**) and body mass index (**BMI**). The size and color of the squares indicate the strength and direction of the correlation. Non- significant genetic correlations are not shown. *r_g_* and 95% confidence intervals can be found in **Supplementary Table 1**. The full genetic correlation matrix is shown in **Supplementary** Figure 1.

### Latent variable models of ED, impulsivity, SUDs and BMI

We first constructed a correlated model of eating disorder phenotypes (AN, AN_Restrictive_, AN_Binge-Purging_, and BE), impulsivity traits, SUDs, and BMI. This model, which is illustrated in **Figure 2**, showed acceptable fit (CFI = 0.94, SRMR = 0.073).

**Figure 2.**
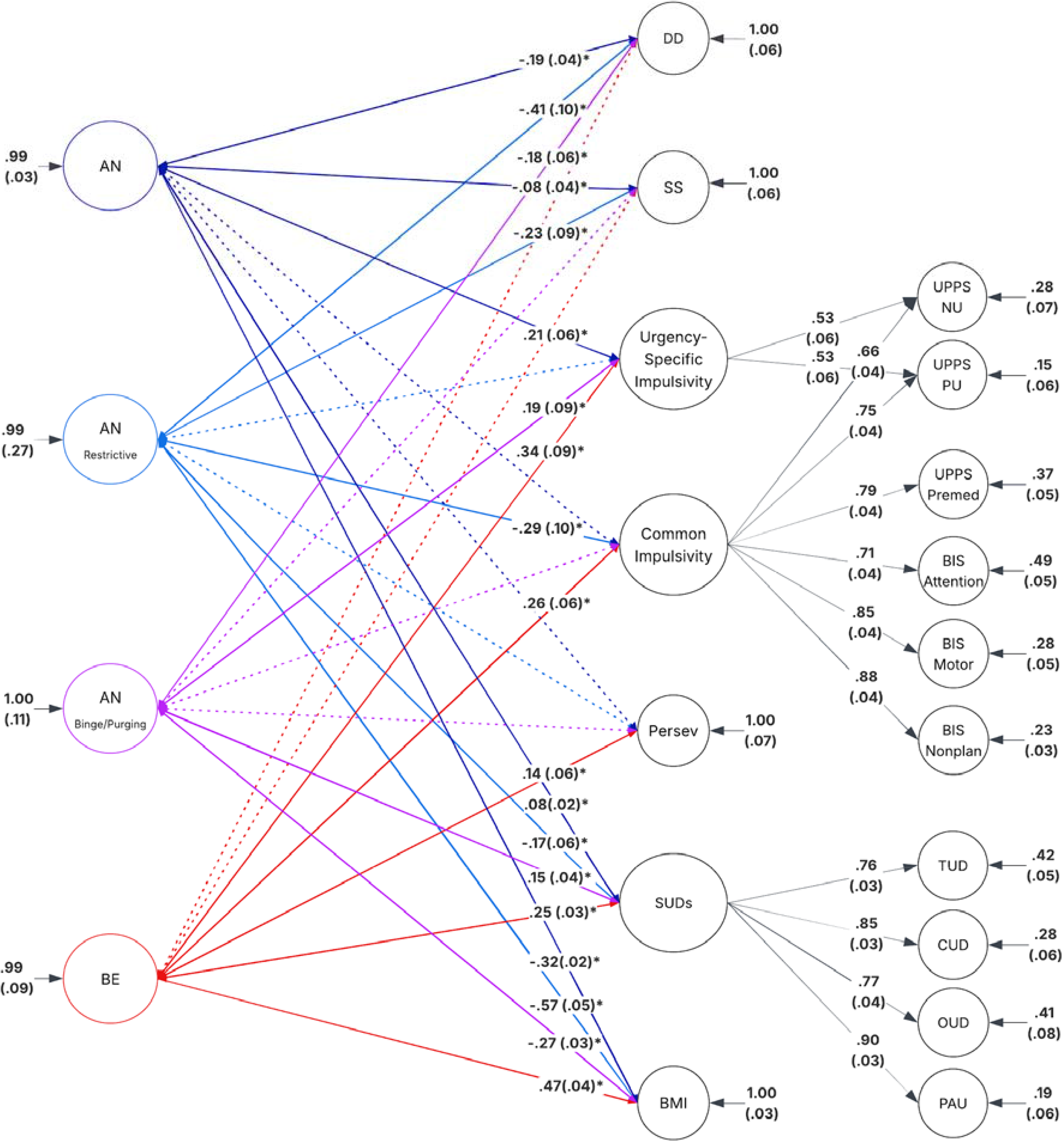
Path diagram for the genetic correlation model with AN, AN_Restrictive_, AN_Binge-Purging_, BE, impulsivity traits, SUDs and BMI. Ovals represent latent factors and squares represent individual observed traits. The “common impulsivity” factor in this model captures the shared variance across selected measures of the BIS and UPPS-P scales. A second latent factor, “urgency-specific impulsivity”, captures the high correlation among UPPS-P negative urgency and UPPS-P positive urgency subscales and is fixed to be uncorrelated with genetic variance in common impulsivity. A single factor, “substance use disorders”, represents the four SUDs. Single headed arrows indicate factor loading, and double headed arrows indicate correlations. Dark blue arrows represent genetic correlations with AN, light blue arrows represent genetic correlations with AN_Restrictive_, purple arrows represent genetic correlations with AN_Binge-Purging_, and red arrows represent genetic correlations with BE. Dotted arrows indicate genetic correlations that were not significant after multiple testing corrections. All values indicate standardized parameter estimates. Standard error and *p* values can be found in **Supplementary Table 2**. UPPS-NU, UPPS-P Negative Urgency; UPPS-PU, UPPS-P Positive Urgency; UPPS-P Premed, UPPS-P Lack of Premediation; BIS Nonplan, BIS Nonplanning; SUDs, substance use disorders; TUD, tobacco use disorder; CUD, cannabis use disorder; OUD, opioid use disorder; PAU, problematic alcohol use; SS, sensation seeking; Persev, Lack of Perseverance; DD, delay discounting; BMI, body mass index; AN, anorexia nervosa; AN_Restrictive_, anorexia nervosa restrictive; AN_Binge-Purging_, anorexia nervosa binge-purging; BE, binge-eating.

AN was negatively genetically correlated with delay discounting (*r_g_* = -0.19 ± 0.04, *p* = 9.26e-07), consistent with our previous study (Bianchi et al., 2025), as well as with sensation seeking (*r_g_* = -0.08 ± 0.04, *p* = 2.95e-02), and positively genetically correlated with urgency- specific impulsivity (*r_g_*= 0.21 ± 0.06, *p* = 9.76e-04).

These genetic correlations showed distinct patterns across AN subtypes. Both AN_Restrictive_ and AN_Binge-Purging_ were negatively genetically correlated with delay discounting, the magnitude of this correlation being significantly stronger for AN_Restrictive_ (*r_g_* = -0.41 ± 0.10, *p* = 6.46e-05) than AN_Binge-Purging_ (*r_g_* = -0.18 ± 0.06, *p* = 5.36e-03; Δχ²(1) = 111.87, *p* = 3.82e-26). AN_Restrictive_ was negatively genetically correlated with sensation seeking (*r_g_* = -0.23 ± 0.09, *p* = 1.17e-02) and common impulsivity (*r_g_* = -0.23 ± 0.09, *p* = 8.99e-03), whereas AN_Binge-Purging_ did not show significant genetic correlations with these facets. Only AN_Binge-Purging_ was positively genetically correlated with urgency-specific impulsivity (*r_g_*= 0.19 ± 0.09, *p* = 4.04e-02).

BE was not significantly genetically correlated with delay discounting or sensation seeking, but was positively genetically correlated with urgency-specific impulsivity (*r_g_* = 0.34 ± 0.09, *p* = 2.94e-4) and common impulsivity (*r_g_* = 0.26 ± 0.06, *p* =7.75e-05). BE was also positively genetically correlated with lack of perseverance (*r_g_*= 0.14 ± 0.06, *p* = 2.83e-02).

All eating disorders showed significant genetic correlations with SUDs, although the magnitude and direction of these estimates varied by subtype. AN and AN_Binge-Purging_ were positively genetically correlated with the SUDs factor (*r_g_*= 0.08 ± 0.02, *p* = 1.78e-03; *r_g_* = 0.15 ± 0.04, *p* = 2.20e-04, respectively) whereas AN_Restrictive_ was negatively genetically correlated (*r_g_*= -0.17 ± 0.06, *p* = 4.55e-03). BE showed the strongest positive *r_g_* with SUDs (*r_g_* = 0.25 ± 0.03, *p* = 2.33e- 13).

AN and both of its subtypes were significantly genetically correlated with BMI: AN_Restrictive_ (*r_g_*= -0.57 ± 0.05, *p* = 5.25e-25), AN (*r_g_* = -0.32 ± 0.02, *p* =4.07e-48), and AN_Binge-Purging_ (*r_g_* = -0.27 ± 0.03, *p* = 4.35e-16). In contrast, BE was positively genetically correlated with BMI (*r_g_* = 0.47 ± 0.04, *p* = 1.87e-30).

We then constructed a multiple regression model, where eating disorder traits were regressed on impulsivity, SUDs, and BMI. This model showed equivalent fit as the correlated factor model, and the results are presented in **Figure 3** and **Supplementary Table 3**. Out of the 19 significant correlations observed in the correlated model, 12 associations remained significant after accounting for shared genetic liability to impulsivity facets, SUDs, and BMI. AN remained negatively associated with delay discounting (β = −0.19 ± 0.08, *p* = 9.24e-03), consistent with our first hypothesis, however, AN_Restrictive_ and AN_Binge-Purging_ no longer showed significant associations with delay discounting. Additionally, both AN and AN_Restrictive_ remained independently negatively associated with sensation seeking (β = -0.11 ± 0.05, *p* = 2.60e-02; β = -0.28 ± 0.13, *p* = 2.64e-02, respectively). While in the correlated model AN_Binge-Purging_ showed a mixed pattern of negative genetic correlations with delay discounting and positive with urgency- related facets, consistent with our second hypothesis, none of these associations were significant in the multiple regression model. BE remained positively associated with urgency- specific impulsivity (β = 0.28 ± 0.12, *p* = 2.21e-02), in line with our third hypothesis. All eating disorder phenotypes remained associated with SUDs and BMI, in the expected direction of effect, with the exception of the association between AN_Restrictive_ and SUDs, which was no longer significant possibly due to limited power.

**Figure 3.**
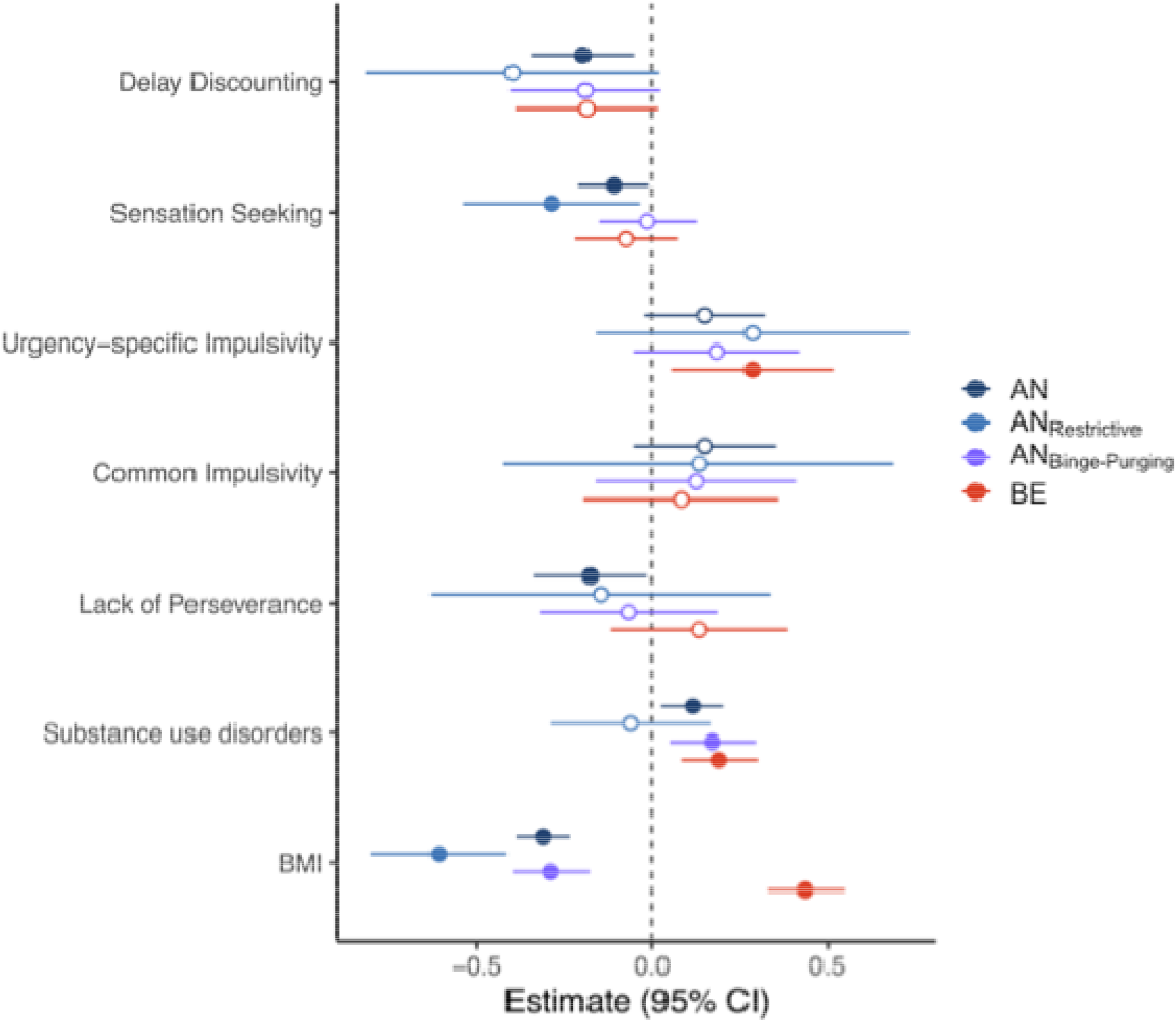
Standardized genetic associations between eating disorder phenotypes — AN, AN_Restrictive_, AN_Binge-Purging_, BE— and impulsivity factors, SUDS, and BMI in a multivariable regression model. Significant associations are indicated by filled-in circles. Estimates, standard error, and *p* values are shown in Supplementary Table 3.

## DISCUSSION

Eating disorders are heterogeneous, and different clinical presentations are characterized by excessive control and/or loss of control over eating. Here, we examined genetic relationships between four eating disorder phenotypes (AN, AN_Restrictive_, AN_Binge-Purging_, and BE) and nine impulsivity phenotypes (five UPPS-P, three BIS-11, and delay discounting), while accounting for shared genetic liability to SUDs and BMI. Our findings provide evidence that (i) eating disorder phenotypes show distinct, sometimes opposing, genetic relationships with specific impulsivity facets; (ii) examining AN subtypes reveals genetic associations that may be obscured when AN is analyzed as a single diagnosis; and (iii) several associations, notably those between AN and delay discounting and sensation seeking, and between BE and urgency-specific impulsivity, persist after accounting for the genetic overlap with BMI and SUDs. These findings suggest that AN cannot be characterized solely by excessive self-control, but may involve a combination of overcontrol and impulsivity that differs across clinical presentations. More broadly, our findings emphasize the importance of examining impulsivity as a multidimensional construct to better understand the genetic heterogeneity of eating disorders and ultimately develop better prevention, diagnosis and treatment strategies.

We previously reported a negative genetic correlation between AN and delay discounting (Bianchi et al., 2025). Here, we replicate this finding using a larger GWAS of AN and extend it to both AN subtypes, with AN_Restrictive_ showing the strongest negative correlation, consistent with our initial hypothesis. These findings mirror phenotypic studies reporting lower delay discounting in patients with AN_Restrictive_ compared to AN_Binge-Purging_ (Amlung et al., 2019; Steinglass et al., 2012; Steward et al., 2017). Lower delay discounting may reflect greater propensity for future-planning (Schuman et al., 2025), potentially contributing to persistent dietary restriction when delayed outcomes, such as thinness and weight loss, are valued over immediate food rewards (Steward et al., 2017). However, this relationship is complex, and additional mechanisms, such as compulsivity, altered reward valuation, and cognitive rigidity, may also contribute to restrictive food intake (Brown et al., 2024; Frost & McNaughton, 2017; Lavender et al., 2017; Peters & Büchel, 2011). For example, some studies suggest that the tendency to delay reward may reflect habitual rather than goal-directed behavior, particularly in individuals with low BMI (Decker et al., 2015; King et al., 2016). In our study, the negative association between AN and delay discounting persisted after accounting for shared genetic liability to BMI, SUDs, and other impulsivity facets, whereas the associations with AN_Restrictive_ and AN_Binge-Purging_ were no longer statistically significant potentially reflecting their smaller sample sizes. Interestingly, delay discounting has been shown to increase following weight restoration in AN (Decker et al., 2015), suggesting that it may be sensitive to changes in illness state.

These findings highlight the potential of delay discounting both as a treatment outcome and a therapeutic target in AN (Amlung et al., 2019; Hoffman et al., 2012; Mallorquí-Bagué et al., 2020; Meneguzzo et al., 2021). While interventions aimed at decreasing delay discounting, such as episodic future thinking, have shown promise in addiction research, developing strategies to increase delay discounting in AN represents a potential avenue for future research (Daniel et al., 2013; Peters & Büchel, 2010; J. S. Stein et al., 2016). Prior studies have suggested that individuals with a decrease in AN symptomatology in recovery have been found to have increased delay discounting (Decker et al., 2015; Lynch et al., 2013; Steinglass et al., 2012).

However, it is possible that the same is not true for other eating disorder phenotypes, such as bulimia nervosa and binge-eating disorder, given their different relationships with impulsivity facets (Amlung et al., 2019; Foldi et al., 2021; Mallorquí-Bagué et al., 2020).

Loss of control is a defining feature of BE, and elevated impulsivity has been associated with binge-eating behaviors in clinical studies (Fernández-Aranda et al., 2008; Meule & Platte, 2015). We observed novel positive genetic correlations between urgency-specific impulsivity and both AN_Binge-Purging_ and BE (Brockmeyer et al., 2014). Although only BE remained significantly associated with this factor in the multivariable regression model, the nonsignificant association for AN_Binge-Purging_ may again reflect power limitations or shared genetic liability with other traits included in the model. Phenotypic studies examining *negative* urgency have found associations between elevated negative urgency and BE episodes in individuals with AN and BE disorder (Culbert et al., 2016; Wonderlich et al., 2024). These findings suggest that BE episodes may serve as a maladaptive response to distress or negative emotions. Consistent with this interpretation, several studies have reported increased negative affect or emotional distress prior to BE episodes, although findings are mixed regarding whether these emotions subside after eating (Borm et al., 2025; Haedt-Matt & Keel, 2011). This suggests that interventions targeting the emotional triggers of BE may be relevant for treating BE and AN_Binge-Purging_ (Yan et al., 2022). In contrast, associations between positive urgency and BE have been less consistent, with some studies reporting that these associations are no longer significant after accounting for negative urgency (Kenny et al., 2019; Yan et al., 2022). In our sensitivity analyses modeling negative and positive urgency separately, neither facet appeared to drive the genetic association with BE on its own (**Supplementary Figure 2**). Given the strong genetic correlation between these two facets, our findings may reflect shared genetic influences on emotion-driven impulsivity rather than effects specific to either negative or positive urgency.

Although prior phenotypic evidence suggests a prominent role for negative urgency, our genetic analyses could not distinguish the contributions of these two facets.

Together, these findings highlight the genetic heterogeneity of eating disorder phenotypes (Termorshuizen et al., 2025). Although AN and its subtypes are strongly genetically correlated, these correlations are below unity, suggesting partly distinct genetic architectures.

The different patterns of genetic overlap with impulsivity broadly mirror differences in their clinical presentations, with AN_Restrictive_ showing stronger negative genetic correlations with delay discounting and common impulsivity, as well as a novel negative genetic correlation with sensation seeking, which is consistent with clinical studies reporting lower novelty seeking and greater avoidance of unfamiliar experiences in individuals with restrictive eating behaviors (Lavender & Mitchell, 2015; Rossier et al., 2000). AN_Binge-Purging_ showed a combination of restrictive and urgency-related features. These contrasting patterns may have implications for understanding transitions between eating disorder phenotypes and diagnoses (Lavender & Mitchell, 2015), including emergence of BE and/or purging behaviors among individuals initially presenting with AN_Restrictive_, or low levels of novelty seeking in individuals with bulimia nervosa who subsequently developed anorexia nervosa (Tozzi et al., 2005). Future longitudinal studies could investigate whether genetic liability to specific impulsivity facets predicts the emergence of BE or purging behaviors, and whether clinical assessment of urgency-specific impulsivity could help identify individuals at risk of developing binge-purging behaviors (Lavender & usMitchell, 2015). Our findings also emphasize the importance of subphenotyping AN to identify genetic associations that may be obscured when analysing AN as a single diagnosis.

Eating disorders share genetic liability with BMI and SUDs, although the direction and magnitude of these relationships vary across phenotypes. For example, AN_Restrictive_ showed a negative genetic correlation with SUDs, whereas AN, AN_Binge-Purging_ and BE showed positive genetic correlations, further illustrating heterogeneity in their relationships with externalizing traits. AN has been conceptualized as a metabo-psychiatric disorder, reflecting genetic influences that extend beyond BMI to other metabolic, lipid, and anthropometric traits (Termorshuizen et al., 2026; Watson et al., 2019). Our results are consistent with this view, showing that several associations persisted after accounting for shared genetic liability to BMI and SUDs, suggesting that these relationships are not fully explained by genetic influences on body mass or substance use. At the same time, the attenuation of other associations suggests that impulsivity, BMI, and SUDs may capture partially overlapping genetic liabilities, highlighting the importance of distinguishing genetic relationships specific to eating disorder phenotypes from those shared with other psychiatric and metabolic traits.

Our findings should be interpreted in the context of several limitations. First, GWAS were restricted to individuals of European-like ancestry, limiting the generalizability of our findings to other ancestry groups. Future studies should extend these analyses to more diverse populations as larger GWAS become available. Second, although Genomic SEM characterizes shared genetic architecture, it cannot establish causality or directionality. Impulsivity facets may represent antecedents or consequences of disordered eating (Boswell et al., 2023; Wonderlich et al., 2024). Longitudinal studies examining changes in eating disorder symptoms alongside impulsivity measures may help clarify these relationships. Third, genome-wide genetic correlations may obscure genomic regions with opposing effects. Complementary methods, such as LAVA and MiXeR, could help identify local genetic correlations and quantify polygenic overlap, respectively, providing further insight into the genetic architecture underlying these associations. Fourth, the GWAS samples for eating disorder phenotypes were predominantly female (96% in BE, 94% in AN), limiting our ability to examine potential sex differences and the generalizability of our findings to males. Finally, the substantially smaller case samples for AN_Restrictive_ and AN_Binge-Purging_ GWAS introduces power imbalances, particularly for detecting associations in the multivariable regression models.

Our work reveals distinct genetic relationships between eating disorder phenotypes and specific impulsivity facets, challenging a simple distinction between excessive control in AN and elevated impulsivity in BE. By distinguishing these relationships across restrictive and binge- eating/purging phenotypes, our study provides a framework for investigating the behavioral and biological mechanisms underlying eating disorder heterogeneity. Ultimately, understanding these shared and distinct genetic influences could inform future prevention and treatment strategies.

## Data Availability

GWAS summary statistics for AN, ANBinge-Purging, ANRestrictive, BE, and the SUD traits are publicly available. Data from 23andMe are available upon request (see https://research.23andme.com/dataset-access/). The R data files containing the genomic SEM matrices for all analyses are displayed at the following link: https://osf.io/6jys3/. This allows for replication and analyses of competing models without obtaining the source data.

## ACKNOWLEDGEMENTS

We would like to thank the research participants and employees of 23andMe for making this work possible.

## FUNDING STATEMENT

This study is supported by the Tobacco-Related Disease Research Program to LVR (award number: T35FT9589), and the National Institutes of Health to MC (award number: T32DA55569, T32AA032229) and SSR (award number: DP1DA054394).

## CONFLICTS OF INTEREST

The authors declare no conflicts of interest.

## Notes

### Competing Interest Statement

The authors have declared no competing interest.

