## Supplementary Figures for "Eating disorder phenotypes differ in their genetic relationships with impulsivity facets"

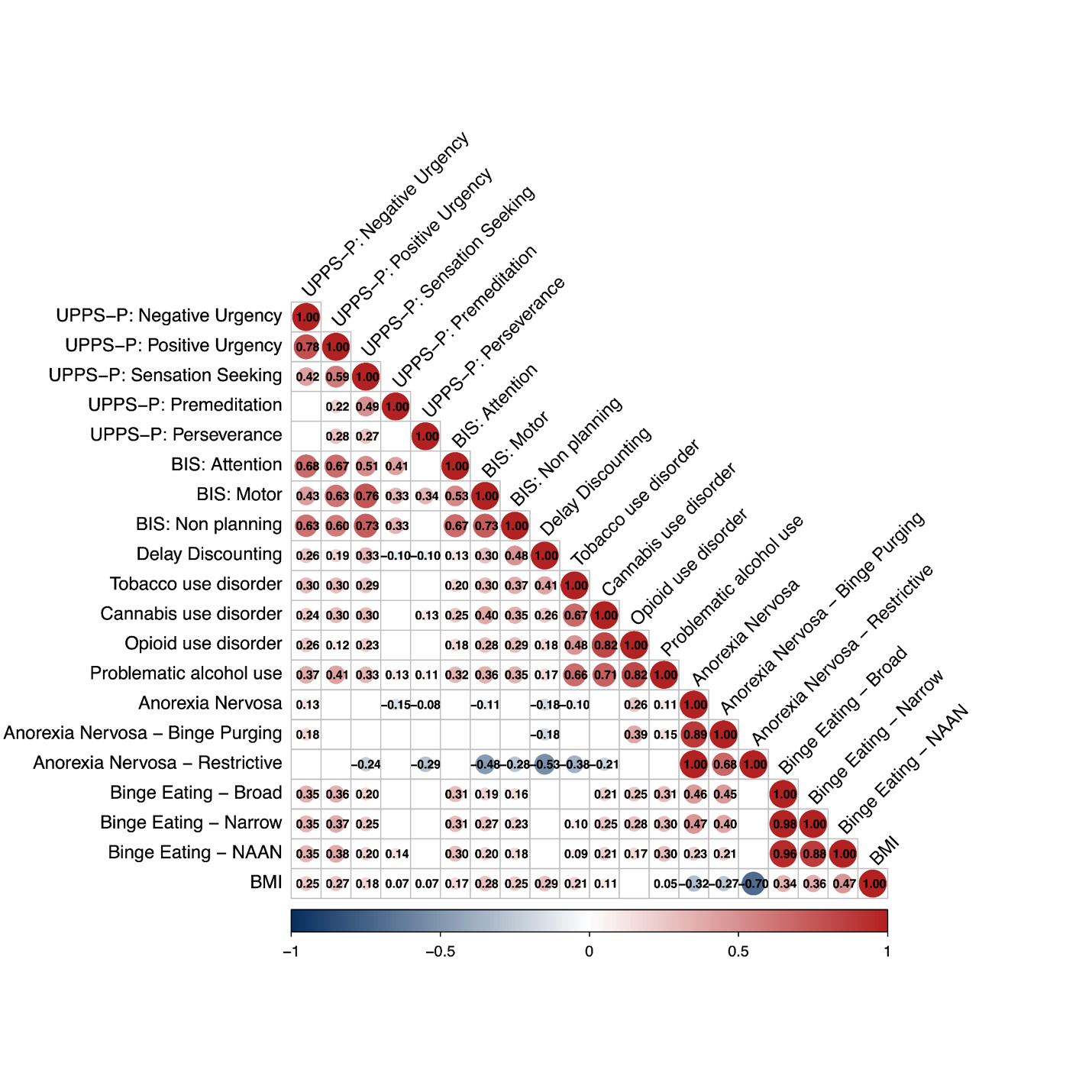

**Supplementary Figure 1.** Genetic correlation matrix among all study variables. In the final study, we included one BE phenotype only, Binge Eating - Non-assertained for AN (NAAN; referred to as BE in the main manuscript), to reduce the number of potentially AN cases in this dataset.

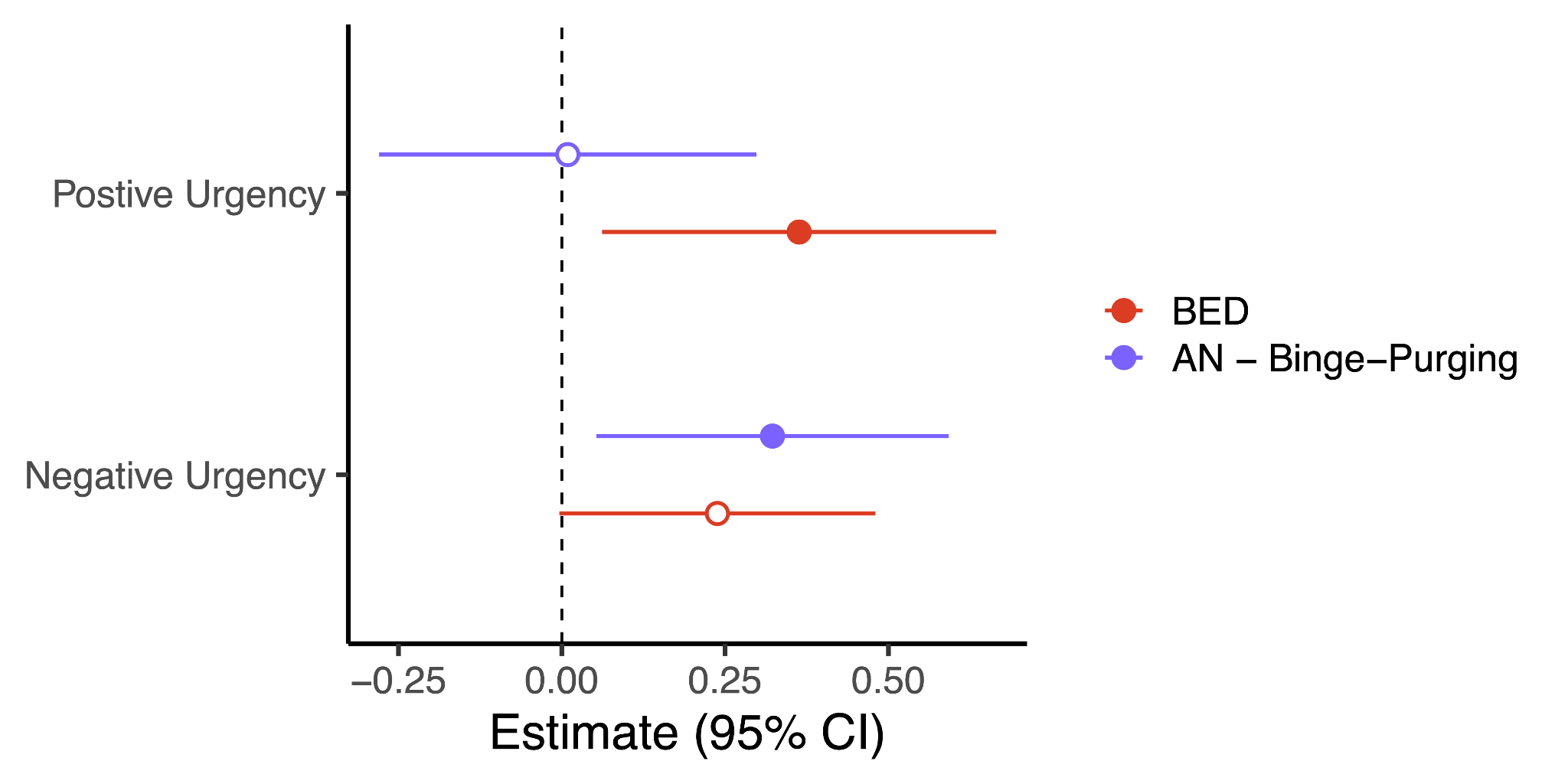

**Supplementary Figure 2.** Sensitivity analysis in the regression model, where positive and negative urgency are modeled separately to test for individual associations with BED and ANBP.
